# Not Sick, Yet Treated: Barriers and Facilitators to Tuberculosis Preventive Treatment Uptake Among Household Contacts of Tuberculosis Patients in Moulvibazar District, Bangladesh

**DOI:** 10.64898/2026.09.28.26364052

**Authors:** Nighat Sultana, Ida Safitri Laksanawati, Mrittika Barua, Ashmita Ghimire, Ichlasul Amalia, Mandira Ajeng Rachmayanthy, Yodi Mahendradhata

## Abstract

**Introduction:** Preventing tuberculosis (TB) through TB Preventive Treatment (TPT) is an essential component of TB control and elimination. In recent years, many countries including Bangladesh, have revised their national programs to extend TPT to household contacts of TB patients. However, coverage remains well below the target, and evidence remains limited on context-specific barriers and facilitators. This study aimed to explore the barriers and facilitators to TPT uptake among household contacts in Bangladesh.

**Methods:** This study followed an exploratory qualitative approach. The setting was Moulvibazar district, Bangladesh. Qualitative data were collected through key-informant interviews with health providers and in-depth interviews with household contacts, and participants were selected through purposive sampling. Thematic analysis was used to explore the factors influencing TPT uptake.

**Results:** Perceived lack of need for treatment among household contacts and side effects of TPT medicines emerged as major barriers. Stigma associated with TB and resource shortage at the health facility level further hampered uptake. Counseling on the importance of TPT and its possible side effects, along with public awareness campaigns, facilitated TPT uptake by addressing household contacts’ concerns. Perceived risk of TB, free TPT services, support from family members, and health providers’ positive attitudes also supported uptake.

**Conclusion:** Addressing household contacts’ concerns about the need for treatment and potential side effects, along with strengthening health facility capacity, is required for improving TPT uptake. Both individual counseling and mass awareness campaigns can help address these concerns and facilitate uptake. These strategies may help increase TPT coverage in Bangladesh and beyond, contributing to global TB elimination efforts.

## Introduction

Tuberculosis (TB) remains one of the world’s most urgent public health challenges. Despite being both preventable and curable, it continues to be the world’s leading cause of death from a single infectious disease (1). More than 10 million new TB cases are reported and more than a million people die from TB each year (2). In addition, an estimated one-quarter of the world’s population is infected with *Mycobacterium tuberculosis*, the bacterium that causes TB (3).

Most people with Latent TB Infection (LTBI) remain asymptomatic, but 5–10% of them will eventually develop active disease in their lifetime (4). Tuberculosis preventive treatment (TPT) can offer 60–90% effectiveness in reducing this risk of progression to active disease in such cases (5). The World Health Organization (WHO) integrated TPT as a cornerstone of the End TB Strategy, particularly for high-risk groups including household contacts of TB patients, people living with HIV, and clinical-risk group people (e.g. on dialysis) (1). TPT options include a weekly dose of isoniazid and rifapentine for 3 months (3HP), or a daily dose of isoniazid and rifampicin for 3 months (3HR), or a daily dose of isoniazid and rifapentine for 1 month (1HP), or a daily dose of rifampicin for 4 months (4R), or a daily dose of isoniazid for 6 or 9 months (6H/9H). Daily 3HR is targeted for infants and young children while weekly 3HP is suitable for adults and children aged above 2 years. 1HP is exclusively reserved for people living with HIV. 6H/9H is suitable for pregnant women or those who cannot tolerate rifamycin. (5).

At the second UN high-level meeting on TB, member states set a target of 90% TPT coverage among household contacts by 2027. By the end of 2024, coverage had reached 25%. This is a substantial gap that must be addressed urgently to meet the global target (6). The shortfall in meeting TPT targets among household contacts highlights the systemic challenges faced by high-burden countries in scaling up TPT (7). Currently, evidence is scarce regarding what factors can improve TPT uptake and what hinders it. Understanding how sociocultural and health system factors interact to influence TPT uptake is also limited, necessitating research to identify context-specific factors that influence uptake (4).

Bangladesh, a lower-middle income country (LMIC) in Southeast Asia, is among the top eight nations that together account for two-thirds of the global TB burden (4, 8). With 221 TB cases per 100,000 population, 25 per 100,000 mortality rates, and an estimated 44 million people with LTBI, the country represents a critical front in the global fight against TB (9). Historically, TB prevention efforts in Bangladesh had focused on children under five (10). In 2021, the National TB Control Program, Bangladesh (NTP) piloted the expansion of TPT to household contacts of all ages in two districts, Moulvibazar and Mymensingh. In 2022, the government decided to scale it up nationwide, marking a major milestone in the country’s TB control strategy (11).

Since the expansion of TPT to household contacts of all ages is a recent initiative both in Bangladesh and globally, understanding the factors affecting its uptake is critical to maximize its impact. This study examined the barriers and facilitators to TPT uptake in Moulvibazar district, Bangladesh. By identifying the influencing factors and outlining actionable strategies to improve uptake, the findings aim to strengthen Bangladesh’s TB control efforts and contribute to achieving the global TPT targets.

## Material and methods

### Study setting

This study took place in Moulvibazar district of Sylhet division, Bangladesh. Moulvibazar was one of the first 2 districts to pilot TPT for household contacts of all ages before the national scale-up. In this district, the TB case notification rate was 253 per 100,000 population against a national rate of 221 per 100,000 population (12). The civil surgeon office (district health office), 2 upazila health complexes (sub-district hospital), Kamalganj and Rajnagar, and 3 Directly Observed Therapy (DOT) centers under these 2 upazilas were selected for data collection. Out of 7 upazilas in Moulvibazar, these 2 upazilas were selected based on the TB case notification rate- one with the highest rate (Kamalganj) and one with the lowest rate (Rajnagar) in the district. Data collection took place between December 2022 to February 2023.

### Study design

This study followed an exploratory qualitative design to explore barriers and facilitators to TPT uptake among household contacts of TB patients.

### Implementation of TPT in Bangladesh

In Bangladesh, TPT for all household contacts was introduced in 2021, and a nationwide scale-up started by 2022. For contacts of bacteriologically confirmed cases, 3HP (once-weekly doses of isoniazid and rifapentine) and 3HR (once-daily doses of isoniazid and rifampin) regimens are in practice. Both regimens require a person to take medicine for 3 months (13).

The Upazila Health Complex (UHC) acts as the primary care point for TB services. Each UHC is equipped with at least one DOT center staffed by NTP and its non-government (NGO) partners. Once a TB patient is diagnosed, health providers at the DOT center identify and assess household contacts either through household visits or by asking them to come to the UHC. Contacts are initially screened for TB symptoms through verbal screening. A physician at the respective UHC then evaluates the contact and recommends tuberculin skin test, chest radiograph, and/or GeneXpert test accordingly. If a person is diagnosed with TB, the physician prescribes anti-TB treatment. If a person is not diagnosed with TB, the physician prescribes TPT. Both medicines can be collected at the DOT center. (13).

Upon treatment initiation, the patient receives a one-month supply of TPT medication to take home. Each month, the patient or a representative can return to collect the next month’s supply. The DOT center conducts monthly follow-ups, either through in-person visits or telephone calls, to ensure treatment adherence.

### Data collection and analysis

Study participants were selected through a purposive sampling approach. We collected qualitative data through 12 key informant interviews (KIIs) among health providers directly involved in TPT implementation and 13 in-depth interviews (IDs) among household contacts. Health providers included both government staff and NGO staff working at the district and sub-district levels. Household contacts included contacts who were either still on TPT or completed TPT within the last 3 months of data collection.

The first author and the research assistant conducted interviews in Bangla using a semi-structured guideline developed based on the TPT care cascade framework, algorithm for TB case finding and preventive treatment, and relevant literature reviews (4, 14). All interviews were recorded using a digital audio recorder and transcribed verbatim. A thematic analysis approach was followed using NVivo qualitative data analysis software (Version 14). Coding was done by the first author in consultation with the co-authors.

### Ethical approval

Ethical approval for this study was obtained from the Ethics Committee of Universitas Gadjah Mada, Yogyakarta, Indonesia (reference no: KE/FK/1320/EC/2022) and Institutional Review Board of BRAC James P. Grant School of Public Health, BRAC University, Bangladesh (reference no: IRB-10 November’22-044).

Written informed consents were obtained for all IDs and KIIs. The information provided by the participants was kept safe, accessible only to the research team, and utilized solely for the purpose of research.

## Results

### Characteristics of study participants

Household contacts’ median age was 36 years (IQR: 25-40 years) and health providers’ median age was 35 years (IQR: 29.5-45.25 years). Health providers included both government and NGO staff working at district-, UHC-, and DOT center-level, and who had at least three months’ experience of providing TB and TPT services. Household contacts included those who were on TPT or completed TPT within the past three months. Table 2 presents the detailed characteristics of participants.

**Table 2.** Characteristics of study participants.

| **Household contacts (N=13)** |  |
| --- | --- |
| Sex | Male (n=5) and female (n=8) |
| Education | Secondary education (n=4), primary education (n=8), none/primary education not completed (n=1) |
| Marital status | Single (n=2), married (n=10), widowed (n=1) |
| TPT status | Completed (n=11), ongoing (n=2) |
| **Health Providers (N=12)** |  |
| Sex | Male (n=10) and female (n=2) |
| Affiliation | Government (n=3) and NGO (N=9) |

### Barriers to TPT uptake

#### Lack of perceived need to take medicines

Household contacts’ lack of perceived need to take medicines emerged as a barrier. Health providers noted that since people with LTBI did not have signs or symptoms of TB, they were reluctant to take tablets. Almost all contacts said that when they first heard about these medicines, they did not understand why they should take tablets as they did not have any disease.

> *“We face one problem over and over again, which is why they (contacts) should take tablets when they are healthy, especially tablets for a serious disease like TB.” (Health provider)*
>
> *“I asked them why I should take medicines when it’s my husband who has TB….They (health providers) told me it’s like corona vaccine, it’s to protect me….then I agreed.” (Household contact)*

#### Adverse side effects of medicines

Side effects from TPT medicines were a barrier, particularly in the continuation phase. Most contacts mentioned experiencing side effects up to some extent, commonly feeling restless or dizzy after taking medicines, nausea, vomiting, stomach pain, red-colored urine, and passing hard stool. Some respondents added that they continued medicines despite these effects while some mentioned skipping doses.

> *“I developed stomach pain in my stomach after starting these medicines. So, I didn’t take them regularly, I started to skip doses….” (Household contact)*

#### Stigma

Some participants expressed concerns about the likelihood of facing negativity from friends, neighbors, or the community because people might assume that since they were taking TB-related medicines, they probably had TB.

> *“People don’t say anything directly, but who knows what’s on their minds! …we used to believe that there’s no cure for TB, it’s scary…people can be afraid” (Household contact)*

#### Shortage of skilled staff

Health providers felt that they were understaffed and it hindered the implementation process and consequently affected uptake. Moreover, with trained employees frequently leaving their jobs, there was a lack of skilled staff.

> *“…we find it difficult to keep track of all the contacts. It hampers the implementation process. We need more staff…” (Health provider)*

#### Infrastructural inadequacy

Health providers mentioned that investigation facilities were often out of order at the UHC level, which caused delays in TPT initiation and eventual dropout. Occasional medicine stock-outs caused further delays. No structured follow-up system was in place for keeping track of the contacts who initiated TPT, which also had negative impacts on the outcome.

> *“Our x-ray machine is out of function now. …We have put the TB patients on treatment already, but we cannot do anything about their family members for the lack of x-ray facilities.” (Health provider)*

### Facilitators to TPT uptake

#### Convincing power of counseling

Counseling on the importance of taking medicines, and prior counseling on the potential drug side effects- what to expect or where to seek help in case of side effects, emerged as a facilitator.

> *“They (contacts) are healthy people, yet they have to take medicines for three months. It’s difficult to do so but it’s possible through counselling.” (Health provider)*
>
> *“I used to feel restless after taking these tablets. I also had red-colored urine. I kept taking tablets because I already knew this would happen, they (health providers) told me earlier.” (Household contact)*

#### Outreach activities

Implementers mentioned that people were concerned about TPT, for example, what it was for or whether it would harm them. Awareness programs and mass campaigns addressed these concerns and had an overall positive impact.

> *“We need to raise awareness on TPT, more people need to know about it. Sometimes we arrange TPT camping (outreach program) at the nearby tea gardens…people appreciate the chance to talk to a doctor. It affects them in a positive way.” (Health provider)*

#### Perceived susceptibility to TB

All household contacts said that TB can spread from one person to another, although they could not always tell the correct mode of transmission. Perceived susceptibility to TB was a facilitator, which encompassed the knowledge that TB can spread. Moreover, watching family members suffer from TB, and the intention to avoid such sufferings in their own lives, helped them to initiate TPT.

> *“…My father-in-law has tuberculosis; what if I get it too? I was scared, so I took tablets.” (Household contact)*

#### Support from family members

Family members’ support influenced individuals on deciding to start TPT. Assurance from the family members, encouragement to keep taking tablets, and support managing side effects played role in both initiation and continuation phase.

> *“I started taking tablets because my mother and my brother told me to. I didn’t think much of it because they would definitely want good for me.” (Household contact)*
>
> *“I consulted with my brother-in-law. …I was afraid that these medicines might harm me. But he told me that it would not cause harm, it was for my own good. So, I decided to start taking tablets.” (Household contact)*

#### Financial assistance to avail of TPT services

TPT medicines are offered free of cost at the primary health center. Transport support is provided for the first time the contacts visit the health center. Participants mentioned this had a positive influence.

> *“I am very happy with the government for providing us with this medicine. I would not be able to buy it with my own money. But now that it is free, my family and I can take it.” (Household contact)*

#### Perceived importance of TPT among health providers

Health providers considered TPT an important part of TB control measures as they believed it had the potential to prevent TB. Some of them expressed concern about but added that they continued to implement TPT as its perceived benefits outweighed the risk.

> *“I think TPT is very important. …it can kill the dormant germs in a person and prevent TB in future. …It’s important for TB control.” (Health provider)*

## Discussion

This study explored barriers and facilitators to TPT uptake among household contacts in Moulvibazar district, Bangladesh. Contacts’ lack of perceived need for TPT and concerns about its side effects emerged as major barriers. Stigma associated with TB and resource shortage at the health facility level further hindered uptake. Counseling of contacts, public awareness campaigns, perceived risk of TB, family encouragement, free TPT services, and health providers’ positive attitudes towards TPT facilitated uptake. Comprehensive counseling and awareness campaigns were particularly impactful in addressing individuals’ concerns about TPT and improving uptake.

In this study, household contacts questioned the need to take medicines for an asymptomatic condition. This finding is consistent with findings from other LMICs, where individuals expressed concern about treating someone who is not “sick”. They questioned the rationale of prescribing medicines for a condition with no noticeable symptoms (14-16). Concerns about TPT side effects further affected treatment initiation and adherence, consistent with evidence that people often discontinue treatment from fear of adverse reactions (17). These findings highlight the need for clear communication about the purpose of TPT; this can help people understand the need for treatment despite not having active disease and address concerns about side effects early.

Stigma surrounding TB was a barrier, discouraging individuals from undergoing screening or initiating TPT. Stigma is rooted in the historical portrayal of TB as a highly contagious and life-threatening disease, and this stigma extends to TPT despite it being a preventive intervention (18, 19). Similar findings have been reported in Indonesia, where stigma prevented caregivers from allowing their children to receive TPT (20). This underscores the importance of shifting the narrative from portraying TB solely as a dangerous disease and reframing it as a preventable condition through right interventions, such as TPT (18, 21).

Health facility-level challenges, such as staff shortages, high turnover, and medicine stock-outs, hampered TPT uptake. Limited diagnostic capacity delayed initial screening, contributing to losses early in the care pathway. These challenges are not unique to Bangladesh, rather a recurring pattern across LMICs (22-25). Although overcoming these barriers requires substantial resource investment, doing so is essential for the long-term success of TPT program.

Findings from this study suggest that both individual counseling and mass awareness approaches could facilitate TPT uptake. Pre-treatment counseling, which explained the purpose of TPT and how to manage potential side effects, was a facilitator. Evidence from other settings has shown that counseling significantly improves treatment adherence by addressing misconceptions and fears (26, 27). Educational initiatives targeting both health workers and the community have been shown to improve acceptance of preventive measures among caregivers of child contacts (26, 28). Being relatively easy to implement and highly impactful, these strategies are particularly well suited for resource-limited settings.

Positive beliefs and attitudes among health workers have been identified as enabler of TPT implementation (17, 29). Health providers in our study demonstrated positive attitudes towards TPT. However, they also expressed concerns about drug resistance and side effects. Although these concerns did not deter them from implementing TPT, these should not be overlooked. Ongoing training and supportive supervision are required to sustain positive attitude.

To the best of our knowledge, this is the first study in Bangladesh which explored the barriers and facilitators to TPT uptake among household contacts. We explored both health providers’ and household contacts’ perspectives, providing a comprehensive picture of the factors influencing uptake. This study has several limitations as well. It was conducted in a single district of Bangladesh, which may limit the generalizability of the findings to other settings. Contacts who refused TPT or were unable to start TPT were not invited for interviews, which may have excluded important insights into barriers.

## Conclusion

This qualitive study explored the barriers and facilitators to TPT uptake among household contacts in Moulvibazar district, Bangladesh. Barriers, including household contacts’ lack of perceived need for TPT, side effects, stigma, and health facility-level challenges, hindered uptake. Addressing these barriers are thus crucial for TPT program success. Facilitators, including counseling household contacts and public awareness campaigns, may be utilized to improve uptake. By identifying these underlying factors, this study provides insights relevant to Bangladesh and other LMICs to inform strategies for increasing TPT coverage as part of global TB elimination efforts.

## Data Availability

The qualitative coding framework, themes, and relevant de-identified data excerpts are available within the paper and its Supporting Information files. The full interview transcripts cannot be shared publicly because they contain sensitive participant information. Data requests for redacted or specific transcript segments can be directed to the Institutional Review Board (IRB) of the BRAC James P Grant School of Public Health, BRAC University, Bangladesh at.

## Acknowledgments

We are grateful to the National Tuberculosis Control Program Office, Sylhet Division, Bangladesh and the Civil Surgeon Office, Moulvibazar, Bangladesh for granting permission to conduct this study. We extend our gratitude to the HEED Bangladesh team for their constant support throughout the study. We thank Pranto Paul for his assistance with fieldwork and data collection. This research was part of a postgraduate thesis undertaken by the first author (NS), and we acknowledge the Special Programme for Research and Training in Tropical Diseases (WHO/TDR) for providing the scholarship that supported the postgraduate degree. Finally, we thank all the participants who generously shared their time and experience with us.

## Author Contributions

Conceptualization: Nighat Sultana

Funding acquisition: Nighat Sultana

Methodology: Nighat Sultana, Ida Safitri Laksanwati, Mrittika Barua, Yodi Mahendradhata

Project administration: Nighat Sultana

Data curation: Nighat Sultana

Formal analysis: Nighat Sultana, Ichlasul Amalia

Validation: Nighat Sultana, Ichlasul Amalia, Mandira Ajeng Rachmayanthy

Supervision: Mrittika Barua, Ida Safitri Laksanwati, Yodi Mahendradhata

Writing – original draft: Nighat Sultana, Ida Safitri Laksanwati, Mrittika Barua, Ashmita Ghimire, Ichlasul Amalia, Mandira Ajeng Rachmayanthy, Yodi Mahendradhata

Writing – review & editing: Nighat Sultana, Ida Safitri Laksanwati, Mrittika Barua, Ashmita Ghimire, Ichlasul Amalia, Mandira Ajeng Rachmayanthy, Yodi Mahendradhata

## Supporting Information

S1 Table. Inclusion and exclusion criteria of study participants

S1 File. Key informant interview and in-depth interview guide

S2 File. Details of codes and themes

