## Supplementary Materials for "Not Sick, Yet Treated: Barriers and Facilitators to Tuberculosis Preventive Treatment Uptake Among Household Contacts of Tuberculosis Patients in Moulvibazar District, Bangladesh"

**Supplementary Table 1. Study participants inclusion and exclusion criteria**

|  | **Inclusion criteria** | **Exclusion criteria** |
| --- | --- | --- |
| Household contacts | - Household contacts of bacteriologically confirmed pulmonary TB cases - Listed in the contact investigation registers maintained by the DOT center in the study area - Aged 19 years and above - Currently receiving 3HP/3HR TPT or completed TPT within the previous three months | - Refuse to participate - Unable to provide informed consent due to cognitive impairment - Individuals with speech or hearing impairment that cannot be reasonably accommodated to complete the interview process |
| Health providers | - Health provider involved in TPT service provision within the study area - Have worked in TPT service provision for three months or more | - Refuse to participate - Individuals with speech or hearing impairment that cannot be reasonably accommodated to complete the interview process |

**Supplementary File 1. Interview guide**

**Interview guide (Bangla)**

**Interview guide for household contacts**

| **প্রধান প্রশ্ন** | **প্রোবস** |
| --- | --- |
| আপনি কি যক্ষারোগ সম্পর্কে জানেন? | 1. যক্ষা সম্পর্কে আপনি কি জানেন?   - রোগের উপসর্গ - কিভাবে ছড়ায় - কার যক্ষা হতে পারে, আপনার বা আপনার পরিবারের কারো হতে পারে কিনা - কিভাবে যক্ষারোগের পরীক্ষা করা হয়, কারা পরীক্ষা করে এবং কোথায় - কিভাবে যক্ষার চিকিৎসা হয়, কতদিন এবং কোথায়   2. আপনি কোথা থেকে এই সকল তথ্য জেনেছেন? |
| আপনার পরিবারের কোন সদস্যের যক্ষারোগ আছে বা অতীতে ছিল? | 1.   - তার সাথে আপনার সম্পর্ক কি? - তার রোগ ধরা পড়েছিল কিভাবে, কোথায় এবং কখন?   2. রোগ ধরা পড়ার পর কি হল-   - সে কি যক্ষার ঔষধ শুরু করেছে? - কতদিন ঔষধ খেয়েছে? - কোথা থেকে ঔষধ সংগ্রহ করত? - ঔষধ খাওয়া শেষ করেছিল? তারপর কি সে যক্ষারোগ মুক্ত হয়েছিল? |
| 2. আপনার পরীবারের সদস্যের যক্ষা হওয়ার পর কি আপনাকে যক্ষারোগের কোন পরীক্ষা করতে বলা হয়েছে? | 1.   - কে পরীক্ষা করতে বলেছে? - কোথায় বলেছে- আপনার বাড়িতে এসে না স্বাস্হ্যকেন্দ্রে না অন্য কোনভাবে? - কি ধরনের পরীক্ষা? কোথায়? - আপনি কি পরীক্ষা করতে গিয়েছেন?   যদি না গিয়ে থাকে, কেন যাননি ?  যদি গিয়ে থাকে,  2. পরীক্ষার ফলাফল কি এসেছিল তা কি আপনি জানেন? এই ফলাফলের মানে কি সে সম্পর্কে আপনার ধারনা আছে?   - পরীক্ষার পর আপনাকে কিছু করতে বলা হয়েছে? - কি করতে বলা হয়েছে (ঔষধ খাওয়া বা অন্য কোন নিয়ম মেনে চলা), কে বলেছে? - আপনি কি তা করতে রাজি হয়েছেন? - আপনি রাজি হবেন কি হবেননা এই সিদ্ধান্তে কিভাবে পৌছালেন? এই সিদ্ধান্ত কি আপনি নিজে নিয়েছেন না অন্য কেউ নিয়েছে বা নিতে সাহায্য করেছে? - ঔষধ খাওয়ার প্রয়োজনীয়তা সম্পর্কে কি আপনাকে বলা হয়েছে? কি বলা হয়েছে? - ঔষধ খেতে হবে এ ব্যাপারে আপনার প্রতিক্রিয়া কি ছিল? - কোথায় ঔষধ পাওয়া যাবে, কখন এবং কিভাবে খেতে হবে, এ ব্যাপারে কি আপনাকে বলা হয়েছে? - ঔষধ খেতে গিয়ে কি আপনাকে কোন ধরনের প্রতিকূলতার সম্মুখীন হতে হয়েছে ? কিরকম ( ঔষধ সংগ্রহ, দূরত্ব, আর্থিক, সামাজিক, কার সাথে ঔষধ আনতে যান ইত্যাদি) ? এ ব্যাপারে কি আপনি কারো সাহায্য চেয়েছেন বা পেয়েছেন (পরিবারের সদস্য বা স্বাস্হ্যকর্মী)? - এভাবে ঔষধ খাওয়ার বিষয়ে আপনার প্রতিক্রিয়া কি?   3. ঔষধের কিছু সম্ভাব্য পার্শ্ব প্রতিক্রিয়া হতে পারে, এ সম্পর্কে আপনাকে বলা হয়েছে?   - কিরকম পার্শ্ব প্রতিক্রিয়া হতে পারে? - এরকম হলে কি করতে হবে বা কোথায় যেতে হবে? - আপনার কি এরকম কোন সমস্যা হয়েছে? - যদি হয়ে থাকে, তখন কি করেছেন- কাকে জানিয়েছেন, কোন সেবা নিয়েছেন? কোথায়? - পরবর্তীতে কি হল- সমস্যার সমাধান হয়েছে? - এরকম পার্শ্ব প্রতিক্রিয়ার ঘটনায় আপনি যে যক্ষাপ্রতিরোধী ঔষধ খাচ্ছেন তার উপর কি প্রভাব পড়েছে?   4. আপনি ঔষধ কতদিন খেয়েছেন (কোর্স শেষ করেছে বা করেনি)?  যদি শেষ না করে থাকে, কেন করেননি?  5. এভাবে ঔষধ খেতে গিয়ে আপনার কোন পর্যায়ে কোন সমস্যার সম্মুখীন হতে হয়েছে? কি ধরনের সমস্যা? এ ব্যাপারে আপনি কারো সাহায্য চেয়েছেন বা পেয়েছেন? কার? |
| যাদের যক্ষা হয়নি কিন্তু ভবিষ্যত যক্ষারোগ প্রতিরোধে ঔষধ খেতে বলা হয়, এই ঔষধের ব্যাপারে আপনার ধারনা কি? | 1. আপনার কি মনে হয় এই ঔষধ খাওয়া দরকার না দরকার নেই? কেন?  যদি ঔষধ খেয়ে থাকে,  2. আপনি যে ঔষধ খাওয়ার সিদ্ধান্ত নিয়েছেন, এর কারন কি ছিল? |
| যাদের যক্ষা নেই কিন্তু যক্ষা প্রতিরোধী ঔষধ খাচ্ছে, সমাজের লোকজন তাদোরকে কিভাবে দেখে? | 1. আপনি যে যক্ষা প্রতিরোধী ঔষধ খাচ্ছেন এটা সমাজের সদস্যদের জানাতে কি স্বাচ্ছন্দ্যবোধ করেন? কেন বা কেন না? |

**Interview guide for DOT center staff**

| **প্রধান প্রশ্ন** | **প্রোবস** |
| --- | --- |
| যক্ষা এবং সুপ্ত যক্ষা বা লেটেন্ট টিবি ইনফেকশন বা এলটিবিআই সম্পর্কে আপনি কি জানেন | 1. যক্ষা নিয়ন্ত্রন প্রোগ্রামে আপনার ভূমিকা কি?   - আপনি কি যক্ষারোগের স্ক্রিনিং করে থাকেন? এটা কিভাবে করা হয়? কোথায়? - যক্ষা রোগ ধরা পড়ে কিভাবে? এর ডায়াগনোসিস কে করে? - এর চিকিৎসা হয় কিভাবে? কোথায়? কতদিন চিকিৎসা করতে হয়?   2. যক্ষা এবং সুপ্ত যক্ষা বা এলটিবিআই এর পার্থক্য সম্পর্কে কি আপনি জানেন?   - এই দুটোর মাঝে কি পার্থক্য রয়েছে? - এলটিবিআই কাদের হতে পারে? এ থেকে পরবর্তীতে কি হতে পারে? - এলটিবিআই এর কোন চিকিৎসা রয়েছে? কি ধরনের চিকিৎসা? কোথায় দেয়া হয়? |
| আপনাকে কি যক্ষা প্রতিরোধী ঔষধ বা টিপিটি নিয়ে কাজ করতে হয়? | 1. আপনাকে কি যক্ষা প্রতিরোধী ঔষধ বা টিপিটি নিয়ে কাজ করতে হয়?   - টিপিটি প্রদানের জন্য কি কি ধাপ রয়েছে? - এই পুরো প্রক্রিয়াটিতে আপনার ভূমিকা কি? - আপনাকে কি কনটাক্ট স্ক্রিনিং করতে হয়? কনটাক্ট স্ক্রিনিং কিভাবে করা হয়? কাদেরকে আপনারা স্ক্রিন করেন (সকল সদস্য না নির্দিষ্ট সংখ্যক সদস্য)? - আপনারা স্ক্রিনিং এর অংশ হিসেবে কি ধরনের প্রশ্ন করেন? কি ধরনের পরীক্ষা করতে বলেন? - আপনারা যাদের কে এসব বলেন তারা এটাকে কিভাবে নেয়- রাজি হয়, তাদেরকে আরো বোঝাতে হয় না প্রত্যাখ্যান করে? - তাদেরকে বোঝাতে গিয়ে আপনার কোনরকম সমস্যার সম্মুখীন হতে হয়? কি ধরনের সমস্যা?   2. একজন কন্টাক্ট টিপিটি পাবে কিনা এই সিদ্ধান্ত কিভাবে নেয়া হয়?   - কে এই সিদ্ধান্ত নেয়? - তারপর কি হয়- কতদিন ঔষধ খেতে হয়? কিভাবে খেতে হয়? কোথায় ঔষধ পাওয়া যায়? - ঔষধ খাওয়া কেন প্রয়োজন এ বিষয়ে কি সেবাগ্রহনকারীকে কাউন্সেলিং করা হয়? কে করে থাকে? - তারা এটাকে কিভাবে নেয়- রাজি হয়, তাদেরকে আরো বোঝাতে হয় না প্রত্যাখ্যান করে?   3. ঔষধের পার্শ্বপ্রতিক্রিয়া সম্পর্কে কি কাউন্সেলিং করা হয়?  কে করে থাকে?   - আপনার কি মনে হয় পার্শ্বপ্রতিক্রিয়ার সম্ভাবনা ঔষধ খাওয়ার সিদ্ধান্তকে প্রভাবিত করে? কিভাবে?   4. আর এমন কোন বিষয় আছে যা টিপিটি দেয়া বা নেয়াকে প্রভাবিত করে?   - কি ধরনের প্রভাব? কিভাবে |
| টিপিটি সম্পর্কে আপনার ধারনা কি ? | 1.   - আপনার কি মনে হয় টিপিটি যক্ষা নিয়ন্ত্রন প্রোগ্রামের একটি গুরুত্বপূর্ন দিক? কেন বা কেন নয়? - টিপিটির এমন কোন দিক আছে যা নিয়ে আপনার সংশয় রয়েছে? কি ধরনের সংশয়? |
| আগে যক্ষা নিয়ন্ত্রন প্রোগ্রাম শুধুমাত্র যক্ষারোগের চিকিৎসা করত, এখন যাদের ভবিষ্যতে যক্ষা হতে পারে তাদেরও চিকিৎসা দেয়া হচ্ছে- এতে প্রোগ্রামে কি ধরনের পরিবর্তন এসেছে? | 1. বর্তমান প্রোগ্রামে টিবি এবং টিপিটি দুটোকেই সমান গুরুত্ব দেয়া হয় না আলাদা?   - কিভাবে? - যদি আলাদা হয়ে, এতে টিপিটির উপর কি ধরনের প্রভাব ফেলে? |
| আপনার ভূমিকা থেকে টিপিটি সম্পর্কিত কাজ করতে গিয়ে আপনি কোন সমস্যার সম্মুখীন হন? | - কি ধরনের সমস্যা? - এই সকল সমস্যা কিভাবে সমাধান করা যায় বা কমানো যায় এ ব্যাপারে আপনার মতামত কি? |

**Interview guide for physicians**

| **প্রধান প্রশ্ন** | **প্রোবস** |
| --- | --- |
| আপনার কাজের অংশ হিসেবে কি টিবি প্রিভেন্টিভ ট্রিটমেন্ট বা টিপিটি নিয়ে কাজ করতে? | 1. টিপিটি ইমপ্লিমেন্টেশনে আপনার ভূমিকা কি?   - কে টিপিটি পাবে বা পাবেনা আপনাকে কি এই সিদ্ধান্ত নিতে হয়? - আপনি কিভাবে এই সিদ্ধান্তে পৌঁছান? এই সিদ্ধান্তে পৌঁছানোর আগে কি কি ধাপ ফলো করতে হয়? - তারপর কি হয়- সেবাগ্রহনকারী বা যে টিপিটি পাবে সে কিভাবে এই সিদ্ধান্তের ব্যপারে জানে, সরাসরি আপনার দ্বারা না স্বাস্হ্যকর্মীর মাধ্যমে? - ঔষধ খাওয়া কেন প্রয়োজন এ বিষয়ে কি সেবাগ্রহনকারীকে কাউন্সেলিং করা হয়? কে করে থাকে?   2. টিপিটির কোন সম্ভাব্য পার্শ্ব প্রতিক্রিয়া রয়েছে?   - ঔষধের পার্শ্বপ্রতিক্রিয়া সম্পর্কে কি কাউন্সেলিং করা হয়? কে করে থাকে? - আপনার কি মনে হয় পার্শ্ব প্রতিক্রিয়ার সম্ভাবনা টিপিটি ইমপ্লিমেন্টেশনকে প্রভাবিত করে? কিভাবে? - পার্শ্ব প্রতিক্রিয়া হয়েছিল এমন কোন কেস পেয়েছেন আপনি? - যদি পেয়ে থাকে, কিরকম পার্শ্ব প্রতিক্রিয়া হয়েছিল? কি চিকিৎসা দিয়েছিলেন? আউটকাম কি ছিল? - আর কোন দিক বা বিষয় আছে যা টিপিটিকে প্রভাবিত করে? কিভাবে? - টিপিটি সাজেস্ট করার সময় কি আপনি রিস্ক-বেনিফিট ব্যালান্স করার চেষ্টা করেন? কিভাবে করেন? |
| টিপিটি সম্পর্কে আপনার ধারনা কি ? | 1.   - আপনার কি মনে হয় টিপিটি যক্ষা নিয়ন্ত্রন প্রোগ্রামের একটি গুরুত্বপূর্ন দিক? কেন বা কেন নয়? - টিপিটির এমন কোন দিক আছে যা নিয়ে আপনার সংশয় রয়েছে? কি ধরনের সংশয়? |
| আপনার ভূমিকা থেকে টিপিটি সম্পর্কিত কাজ করতে গিয়ে আপনি কোন সমস্যার সম্মুখীন হন? | 1.   - কি ধরনের সমস্যা? - এই সকল সমস্যা কিভাবে সমাধান করা যায় বা কমানো যায় এ ব্যাপারে আপনার মতামত কি? |

**Interview guide for district TB program manager**

| **প্রধান প্রশ্ন** | **প্রোবস** |
| --- | --- |
| আপনার কাজের অংশ হিসেবে কি টিবি প্রিভেন্টিভ ট্রিটমেন্ট বা টিপিটি নিয়ে কাজ করতে? | 1.   - টিপিটি ইমপ্লিমেন্টেশনে আপনার ভূমিকা কি? - টিপিটি ইমপ্লিমেন্টেশনে কি কি ধাপ রয়েছে? - কনটাক্ট স্ক্রিনিং কারা করে থাকে? কিভাবে করা হয়? কাদেরকে স্ক্রিন করা হয়? - কি ধরনের প্রশ্ন করা হয়? কি ধরনের পরীক্ষা করতে বলা হয় - তাদেরকে বোঝাতে গিয়ে কর্মীরা কোন সমস্যার সম্মুখীন হয়? কি ধরনের সমস্যা?   2. একজন কন্টাক্ট টিপিটি পাবে কিনা এই সিদ্ধান্ত কিভাবে নেয়া হয়?   - কে এই সিদ্ধান্ত নেয়? - তারপর কি হয়- কতদিন ঔষধ খেতে হয়? কিভাবে খেতে হয়? কোথায় ঔষধ পাওয়া যায়? - ঔষধ খাওয়া কেন প্রয়োজন এ বিষয়ে কি সেবাগ্রহনকারীকে কাউন্সেলিং করা হয়? কে করে থাকে? - তারা এটাকে কিভাবে নেয়- রাজি হয়, তাদেরকে আরো বোঝাতে হয় না প্রত্যাখ্যান করে?   2. টিপিটির কোন সম্ভাব্য পার্শ্ব প্রতিক্রিয়া রয়েছে?   - ঔষধের পার্শ্বপ্রতিক্রিয়া সম্পর্কে কি কাউন্সেলিং করা হয়? কে করে থাকে? - আপনার কি মনে হয় পার্শ্ব প্রতিক্রিয়ার সম্ভাবনা টিপিটি ইমপ্লিমেন্টেশনকে প্রভাবিত করে? কিভাবে?   3.আর কোন দিক বা বিষয় আছে যা টিপিটিকে প্রভাবিত করে? কিভাবে? |
| টিপিটি সম্পর্কে আপনার ধারনা কি ? | 1.   - আপনার কি মনে হয় টিপিটি যক্ষা নিয়ন্ত্রন প্রোগ্রামের একটি গুরুত্বপূর্ন দিক? কেন বা কেন নয়? - টিপিটির এমন কোন দিক আছে যা নিয়ে আপনার সংশয় রয়েছে? কি ধরনের সংশয়? |
| আগে যক্ষা নিয়ন্ত্রন প্রোগ্রাম শুধুমাত্র যক্ষারোগের চিকিৎসা করত, এখন যাদের ভবিষ্যতে যক্ষা হতে পারে তাদেরও চিকিৎসা দেয়া হচ্ছে- এতে প্রোগ্রামে কি ধরনের পরিবর্তন এসেছে? | 1. বর্তমান প্রোগ্রামে টিবি এবং টিপিটি দুটোকেই সমান গুরুত্ব দেয়া হয় না আলাদা?   - কিভাবে? - যদি আলাদা হয়ে, এতে টিপিটির উপর কি ধরনের প্রভাব ফেলে? |
| টিপিটি ইমপ্লিমেন্টেশনে কি কি চ্যালেন্জ রয়েছে? | 1.   - এই সকল চ্যালেন্জ কিভাবে সমাধান করা যায় বা কমানো যায় এ ব্যাপারে আপনার মতামত কি? |

**Interview guide for TB and Leprosy Control Assistant (NTP) / TB Control Assistant (NGO)**

| **প্রধান প্রশ্ন** | **প্রোবস** |
| --- | --- |
| যক্ষা এবং সুপ্ত যক্ষা বা লেটেন্ট টিবি ইনফেকশন বা এলটিবিআই সম্পর্কে আপনি কি জানেন | 1. যক্ষা নিয়ন্ত্রন প্রোগ্রামে আপনার ভূমিকা কি?   - আপনি কি যক্ষারোগের স্ক্রিনিং করে থাকেন? এটা কিভাবে করা হয়? কোথায়? - যক্ষা রোগ ধরা পড়ে কিভাবে? এটার ডায়াগনোসিস কে করে? - এর চিকিৎসা হয় কিভাবে? কোথায়? কতদিন চিকিৎসা করতে হয়?   2. যক্ষা এবং সুপ্ত যক্ষা বা এলটিবিআই এর পার্থক্য সম্পর্কে কি আপনি জানেন?   - এই দুটোর মাঝে কি পার্থক্য রয়েছে? - এলটিবিআই কাদের হতে পারে? এ থেকে পরবর্তীতে কি হতে পারে? - এলটিবিআই এর কোন চিকিৎসা রয়েছে? কি ধরনের চিকিৎসা? কোথায় দেয়া হয়? |
| আপনাকে কি যক্ষা প্রতিরোধী ঔষধ বা টিপিটি নিয়ে কাজ করতে হয়? | 1. আপনাকে কি যক্ষা প্রতিরোধী ঔষধ বা টিপিটি নিয়ে কাজ করতে হয়?   - টিপিটি প্রদানের জন্য কি কি ধাপ রয়েছে? - এই পুরো প্রক্রিয়াটিতে আপনার ভূমিকা কি? - আপনাকে কি কনটাক্ট স্ক্রিনিং করতে হয়? কনটাক্ট স্ক্রিনিং কিভাবে করা হয়? কাদেরকে আপনারা স্ক্রিন করেন (সকল সদস্য না নির্দিষ্ট সংখ্যক সদস্য)? - আপনারা স্ক্রিনিং এর অংশ হিসেবে কি ধরনের প্রশ্ন করেন? কি ধরনের পরীক্ষা করতে বলেন? - আপনারা যাদের কে এসব বলেন তারা এটাকে কিভাবে নেয়- রাজি হয়, তাদেরকে আরো বোঝাতে হয় না প্রত্যাখ্যান করে? - তাদেরকে বোঝাতে গিয়ে আপনার কোনরকম সমস্যার সম্মুখীন হতে হয়? কি ধরনের সমস্যা?   2. একজন কন্টাক্ট টিপিটি পাবে কিনা এই সিদ্ধান্ত কিভাবে নেয়া হয়?   - কে এই সিদ্ধান্ত নেয়? - তারপর কি হয়- কতদিন ঔষধ খেতে হয়? কিভাবে খেতে হয়? কোথায় ঔষধ পাওয়া যায়? - ঔষধ খাওয়া কেন প্রয়োজন এ বিষয়ে কি সেবাগ্রহনকারীকে কাউন্সেলিং করা হয়? কে করে থাকে? - তারা এটাকে কিভাবে নেয়- রাজি হয়, তাদেরকে আরো বোঝাতে হয় না প্রত্যাখ্যান করে?   3. ঔষধের পার্শ্বপ্রতিক্রিয়া সম্পর্কে কি কাউন্সেলিং করা হয়? কে করে থাকে?  আপনার কি মনে হয় পার্শ্বপ্রতিক্রিয়ার সম্ভাবনা ঔষধ খাওয়ার সিদ্ধান্তকে প্রভাবিত করে? কিভাবে?  4. আর এমন কোন বিষয় আছে যা টিপিটি দেয়া বা নেয়াকে প্রভাবিত করে?   - কি ধরনের প্রভাব? কিভাবে |
| টিপিটি সম্পর্কে আপনার ধারনা কি ? | 1.   - আপনার কি মনে হয় টিপিটি যক্ষা নিয়ন্ত্রন প্রোগ্রামের একটি গুরুত্বপূর্ন দিক? কেন বা কেন নয়? - টিপিটির এমন কোন দিক আছে যা নিয়ে আপনার সংশয় রয়েছে? কি ধরনের সংশয়? |
| আগে যক্ষা নিয়ন্ত্রন প্রোগ্রাম শুধুমাত্র যক্ষারোগের চিকিৎসা করত, এখন যাদের ভবিষ্যতে যক্ষা হতে পারে তাদেরও চিকিৎসা দেয়া হচ্ছে- এতে প্রোগ্রামে কি ধরনের পরিবর্তন এসেছে? | 1. বর্তমান প্রোগ্রামে টিবি এবং টিপিটি দুটোকেই সমান গুরুত্ব দেয়া হয় না আলাদা?   - কিভাবে? - যদি আলাদা হয়ে, এতে টিপিটির উপর কি ধরনের প্রভাব ফেলে? |
| আপনার ভূমিকা থেকে টিপিটি সম্পর্কিত কাজ করতে গিয়ে আপনি কোন সমস্যার সম্মুখীন হন? | 1.   - কি ধরনের সমস্যা? - এই সকল সমস্যা কিভাবে সমাধান করা যায় বা কমানো যায় এ ব্যাপারে আপনার মতামত কি? |

**Interview guide (English)**

**Interview guide for household contacts**

| **Main question** | **Probes** |
| --- | --- |
| Do you know about TB? | 1. What do you know about TB   - sign symptoms - mode of transmission - who can get infected and whether family members can get infected - how it is screened, who does the screening and where - how it is treated, for how long and where   2. From where you know about this information? |
| Does anyone in your family have TB or had TB in the past? | 1. Who is that person (relationship with the participant)?  2. How was she or he diagnosed with TB (how, when and where)?  3. What happened after diagnosis,   - did they start taking anti TB medications? - For how long did they take medications? - Where did they get the medications from? - What was the final outcome (was treatment completed, were they cured)? |
| Were you given any instructions to test for TB after your family member got diagnosed with TB? | 1. What instructions were you given   - by whom? - where- home or health care centre? - what kind of tests were advised? - Did you go for the tests? - If no why not?   If yes, what was the test result? Did you know what that result mean?  3. What did you do after the test,   - did you receive any instructions after the test? - what instructions (medications or other preventive measures) and from whom? - Did you follow the instructions? - How did you come to that decision of whether to follow or not follow the instructions? who took that decision? - Were you informed about why you need to take medications? - how did you feel about taking medications? - did you know where to get the medications and when or how to take it? - Did you face any challenge while taking medications (what kind of challenges- accessing medications, distance, financial, social, stigma, who does she or he go with), did you seek or get help from health workers to overcome those challenges? - how did you feel about taking medications this way?   4. Were you informed about the possible adverse events related to medications?   - what kind of adverse events - where to go in case of adverse events - did you experience any adverse events - if yes, did you seek any care after that- where and how - what was the outcome- resolved or not, did it affect your medication status?   5. For how long did you take medications (completed or not)?  If not completed, why not?  6. Did you face any problem at any point to follow the instruction or taking medications? What kind of difficulties and why? Did you seek any help to overcome those difficulties? From who (family members or health care workers)  7. Were any of your family members given any instructions to test or take medications the same as you? Did they follow the instructions? How did they come to take that decision, who made the decision? |
| What is your perception of taking medications to prevent future TB? | 1. Do you think taking medications to prevent future TB is important? Why or why not?  If the person took medications,  2. What motivated you to take medications? |
| How do you think people in your community perceive people who does not have TB but take medications to prevent future TB? | 1. Are you comfortable in disclosing your medication status to other people? Why or why not? |

**Interview guide for DOT center staff**

| **Main question** | **Probes** |
| --- | --- |
| What do you know about TB and latent TB infection or LTBI? | 1.What are your roles and responsibilities in the TB control programme?   - How do you screen for TB? Who does the screening? Where? How (can you explain the procedure?) - How is it diagnosed? By whom? - How is it treated? Where is the treatment available? What is the treatment duration?   2. Do you know the difference between TB and LTBI?   - How are they different? - Who can get LTBI? - What happens after LTBI? - Is treatment available? what kind of treatment? - where is the treatment available? |
| Does your job involve TPT implementation? | 1. Can you tell us about the steps of TPT implementation?   - what is your role in this implementation process? - Do you have to conduct contact screening? - Who are the contacts? how do you do contact screening? Who do you screen (all members or some)? - What kind of questions do you ask? What kind tests do you advise? - How do people respond when you ask those questions or tell them to come for tests (agree, need convincing or refuse)? - Do you face any challenge convincing them? What kind of challenges?   2. Whether a person is eligible for TPT or not- how is that decision taken?   - Who take that decision? - What happens after that? - For how long they need to take medications? How frequently? - Where do they take the medication from? - Do beneficiaries get counselling about the need for taking TPT? Who counsel them? - how do beneficiaries respond to the advice of taking TPT (agree, refuse or need more convincing)?   3. Are beneficiaries informed and counselled about adverse events prior to treatment start? By whom?   - Do you think the possibility of adverse events affect TPT implementation or its utilization? How?   4. Do you think any other factor affect TPT implementations?   - what other factors - what kind of effect, and how? |
| What is your perception of TPT? | - Do you think treating LTBI or TPT is an important part of TB control programme? Why or why not? - Do you have any doubts about the need for TPT? What kind of doubts? |
| TB control program used to treat active TB earlier, now that TPT is added, do you think the program focus has changed? | 1. Does the TB control program give same priority to treatment and preventive treatment or is it different?   - If different, how is it different? - Does that affect TPT implementation? How? |
| In your role, do you face any challenge implementing TPT? | - What kind of challenges do you face? - What do you think can be done to mitigate or overcome those challenges? |

**Interview guide for physicians**

| **Main question** | **Probes** |
| --- | --- |
| Can you tell us about TPT? | 1. In your job, do you evaluate people for TPT? How?   - What is your role in this implementation process? - Whether a person is eligible for TPT or not-Do you have to take the decision? How do you come to the decision? - What steps are followed to reach that decision? - What happens after that- how do beneficiaries get the information and by whom (directly from the physician or from health care workers)? - Are they counselled about the need for taking TPT? By whom? - How do they respond to the decision (agree to take medications, need convincing or refuse)?   2. Are there possible adverse events when suggesting TPT? What are they?   - Do you think the possibility of adverse events affect TPT implementation? How? - Did you have to treat any case of adverse events? - If yes, what were the events? How did you treat them? What was the outcome? - Are the beneficiaries counselled about adverse events? By whom? - Do you try to maintain the risk-benefit balance when suggesting TPT? How do you do that? |
| What is your perception of TPT? | - Do you consider TPT as an important component of TB control programme? Why or why not? - Do you have any doubts about the need for TPT? What kind of doubts? |
| In your role, do you face any challenge implementing TPT? | - What kind of challenges do you face? - What do you think can be done to mitigate or overcome those challenges? |

**Interview guide for district TB program manager**

| **Main question** | **Probes** |
| --- | --- |
| Can you tell us about TPT? | 1. Does your job involve TPT implementation?   - What are the steps of TPT implementation? - What is your role in this implementation process? - Who conduct contact screening? How is it done? Who do you screen (all members or some)? - What kind of questions are asked? What kind tests are advised? - How do people respond to those questions or advice for tests (agree, need convincing or refuse)? - Are there any challenges convincing them? What kind of challenges?   2. Whether a person is eligible for TPT or not- how is that decision taken?   - Who take that decision? - What happens after that? - For how long they need to take medications? How frequently? - Where do they take the medication from? - Do beneficiaries get counselled about the need for taking TPT? Who counsel them? - how do beneficiaries respond to the advice of taking TPT (agree, refuse or need more convincing)?   3. Are there possibility of adverse events if a person takes TPT?   - Are beneficiaries informed and counselled about adverse events prior to treatment start? By whom? - Do you think the possibility of adverse events affect TPT implementation or its utilization? How?   4. Do you think any other factor affect TPT implementations?   - what other factors - what kind of effect, and how? |
| What is your perception of TPT? | - Do you consider TPT as an important component of TB control programme? Why or why not? - Do you have any doubts about the need for TPT? What kind of doubts? |
| TB control program used to treat active TB earlier, now that TPT is added, do you think the program focus has changed? | 1. Does the TB control program give same priority to treatment and preventive treatment or is it different?   - If different, how is it different? - Does that affect TPT implementation? How? |
| Are there challenges to implementing TPT? | - What kind of challenges are there? - What do you think can be done to mitigate or overcome those challenges? |

**Interview guide for TB and Leprosy Control Assistant (NTP) / TB Control Assistant (NGO)**

| **Main question** | **Probes** |
| --- | --- |
| What do you know about TB and latent TB infection or LTBI? | 1.What do you know about TB?   - How is it screened? Who does the screening? - How is it diagnosed? - How is it treated? Where is the treatment available?   2. Do you know the difference between TB and LTBI?   - How are they different? - Who can get LTBI? - What happens after LTBI? - Is treatment available? what kind of treatment? - where is the treatment available? |
| Does your job involve TPT implementation? | 1. Can you tell us about the steps of TPT implementation?   - What is your role in this implementation process? - Who conduct contact screening? How is it done? Who do you screen (all members or some)? - What kind of questions are asked? What kind tests are advised? - How do people respond to those questions or advice for tests (agree, need convincing or refuse)? - Are there any challenges convincing them? What kind of challenges?   2. Whether a person is eligible for TPT or not- how is that decision taken?   - Who take that decision? - What happens after that? - For how long they need to take medications? How frequently? - Where do they take the medication from? - Do beneficiaries get counselled about the need for taking TPT? Who counsel them? - how do beneficiaries respond to the advice of taking TPT (agree, refuse or need more convincing)?   3. Are there possibility of adverse events if a person takes TPT?   - Are beneficiaries informed and counselled about adverse events prior to treatment start? By whom? - Do you think the possibility of adverse events affect TPT implementation or its utilization? How?   4. Do you think any other factor affect TPT implementations?   - what other factors - what kind of effect, and how? |
| What is your perception of TPT? | - Do you think treating LTBI or TPT is an important part of TB control programme? Why or why not? - Do you have any doubts about the need for TPT? What kind of doubts? |
| TB control program used to treat active TB earlier, now that TPT is added, do you think the program focus has changed? | 1. Does the TB control program give same priority to treatment and preventive treatment or is it different?   - If different, how is it different? - Does that affect TPT implementation? How? |
| Are there challenges to implementing TPT? | - What kind of challenges are there? - What do you think can be done to mitigate or overcome those challenges? |

**Supplementary File 2. Details of codes and themes**

**Codes and themes with example quotes**

| **Code** | **Theme** | **Example quote** |
| --- | --- | --- |
| Perception about being healthy | Lack of perceived need to take medicines | “*Some people refuse to start or continue TPT. It’s understandable. Even sick people don’t like taking medications. Here we are telling apparently healthy people to take medications for quite a long time, it’s hard to convince them.*” Health provider |
| Medicine for ‘no disease’ |  |  |
| Feeling unwell | Adverse side effects of medicines | “*I developed pain in my stomach after starting these medicines. So, I didn’t take them regularly, I started to skip doses….”* Household contact |
| Needed to see a doctor |  |  |
| Had to take medicine |  |  |
| Community attitude towards TB patients | Stigma | “*They know about my wife getting treated for TB, but they don’t know that we take medications too. If they knew, they would take it negatively. They would think that we have TB too and would stay away from us. People fear the disease.*” Household contact  “*Our neighbors know that we all take medications. They have no issue with it.*” Household contact |
| Community understanding about preventive treatment |  |  |
| Willingness to reveal TPT status outside the family |  |  |
| Expected outcome of sharing TPT status to others |  |  |
| Frequent staff turnover | Shortage of skilled staff | “*We find it difficult to keep track of all TPT-eligible contacts. Sometimes we visit their homes, sometimes we reach them over phone. But our coverage area is quite big, and we do not have that many people in our center. It hampers the implementation process. We need more staff...*” Health provider |
| Lack of training |  |  |
| Staff shortage |  |  |
| Lack of investigation facilities | Infrastructural inadequacy | “*Our x-ray machine is out of function now. ...We have put the TB patients on treatment already, but we cannot do anything about their family members for the lack of x-ray facilities.”* Health provider |
| Unavailability of medicines |  |  |
| Lack of a structured follow-up system |  |  |
| Counseling on the purpose of TPT | Convincing power of counseling | “*Counseling is the most important part here. TPT-eligible contacts are not patients, they are healthy people, yet they have to take medications for three months. It’s difficult to convince them to do so but it’s possible through counseling. They understand if we use the right approach. Compared to the past, TPT outcome is better now and it’s because of counseling.”* Health provider |
| Prior counseling on the adverse side effects |  |  |
| Involving influential people in the community |  |  |
| Organizing TPT camps (tea garden centered awareness program) | Outreach activities | *“We need to raise awareness on TPT, more people need to know about it. Sometimes we arrange TPT camps (outreach program) at the nearby tea gardens...people appreciate the chance to talk to a doctor. It affects them in a positive way.*” Health provider |
| Mass awareness campaigns |  |  |
| TB transmission | Perceived susceptibility to TB | *“…My father-in-law has tuberculosis; what if I get it too? I was scared, so I took tablets.”* Household contact |
| Perceived risk of TB |  |  |
| Watching family members suffer |  |  |
| Encouragement from family members | Support from family members | “*I work in a mosque here. Sometimes I cannot come to the health center because of my work… my father came to collect my medications.”* Household contact |
| Flexibility of medicine collection |  |  |
| Free of cost medicines | Financial assistance to avail of TPT services | “*I am very happy with the government for providing us with this medicine. I would not be able to buy it with my own money. But now that it is free, my family and I can take it.”* Household contact |
| Transport support |  |  |
| Health providers home-delivering medicines |  |  |
| Perceived effectiveness of TPT | Perceived importance of TPT among health providers | *“I think TPT is very important. …it can kill the dormant germs in a person and prevent TB in future. … It’s important for TB control.”* Health provider |
| Concerns about drug side effects |  |  |
| Concerns about drug resistance |  |  |
| TPT implementation measures |  |  |

**Codebook**

| **Codes** | **Definition** |
| --- | --- |
| Community attitude towards TB patients | Respondents described whether TB patients face any negative attitude in the community they live in |
| Community understanding about preventive treatment | Respondents described whether people understood the difference between treatment for active TB and preventive treatment |
| Concerns about drug resistance | Respondents described whether they were concerned about the possibility of TPT medicines causing TB drug resistance and how it affected their decision to implement TPT |
| Concerns about drug side effects | Respondents described whether they were concerned about the possibility of TPT medicines causing side effects and how it affected their decision to implement TPT |
| Counseling on the purpose of TPT | Respondents described how household contacts are approached before initiating TPT, whether they were informed about the need to take TPT medicines and how it affected decision making |
| Encouragement from family members | Respondents described their family members’ influence on initiating or continuing TPT |
| Expected outcome of sharing TPT status to others | Respondents described their thoughts on what might happen if they shared with their friends or neighbors about taking TPT, whether they would take it positively or negatively |
| Feeling unwell | Respondents described the sense of not feeling well or experiencing undesirable effects after taking TPT medicines |
| Flexibility of medicine collection | Respondents described their family members collecting medicines on their behalf and how it helped them to continue TPT |
| Free of cost medicines | Respondents described that they TPT medicines were available free of cost and how it affected their decision to take TPT |
| Frequent staff turnover | Respondents described experienced employees frequently leaving jobs and those roles getting replaced by new employees who needed time to build program skills |
| Health providers home-delivering medicines |  |
| Involving influential people in the community | Respondents described community leaders’ and religious leaders’ roles in convincing people to take TPT |
| Lack of a structured follow-up system | Respondents described not having a formal or structured follow-up system to track compliance over 3 months regimen |
| Lack of investigation facilities | Respondents described investigation facilities required for starting TPT, such as chest radiograph and tuberculin skin test, not being available at the health center |
| Lack of training | Respondents described whether they underwent training on TPT and whether they felt confident about their roles in TPT implementation |
| Mass awareness campaigns | Respondents described how large-scale community awareness programs influenced TPT implementation |
| Medicine for ‘no disease’ | Respondents described what they thought about taking medicines even though they did not have any disease |
| Had to take medicine | Respondents described experiencing undesirable effects related to TPT which led them to take medicines to relieve those symptoms |
| Needed to see a doctor | Respondents described experiencing undesirable effects related to TPT which led them consult a doctor |
| Organizing TPT camps (tea garden centered awareness program) | Respondents described organizing tea garden centered TPT programs for mass contact screening and its impact on TPT outcome |
| Perceived effectiveness of TPT | Respondents described whether they considered TPT as effective or necessary for controlling TB |
| Perceived risk of TB | Respondents described whether they considered themselves being at risk of being infected with TB or not |
| Perception about being healthy | Respondents described whether they perceived them as healthy and what they meant by it, particularly in relation to TB |
| Prior counseling on the adverse side effects | Respondents described whether they were told about the possibility of adverse side effects of medication before starting TPT and how it affected their decision making |
| Staff shortage | Respondents described having or not having enough staff compared to the workload |
| TB transmission | Respondents described their understanding whether TB can spread from person to person and if yes, how |
| TPT implementation measures | Respondents described their role in TPT implementation and what measures they followed to ensure implementation |
| Transport support | Respondents described getting free of cost transport to visit health care centers for screening and TPT medicine collection |
| Unavailability of medicines | Respondents described all TPT regimen not being available all the time. For example, only 3HP being available at a time or only 3HR being available at a time |
| Watching family members suffer | Respondents described watching their family members being diagnosed with TB and how it affected them |
| Willingness to reveal TPT status outside the family | Respondents described whether they felt comfortable sharing that they were taking TPT with their friends or neighbors |
